# GENOMIC CHARACTERISATION AND PHYLOGENETIC ANALYSIS OF SARS-COV-2 IN ITALY

**DOI:** 10.1101/2020.03.15.20032870

**Authors:** Gianguglielmo Zehender, Alessia Lai, Annalisa Bergna, Luca Meroni, Agostino Riva, Claudia Balotta, Maciej Tarkowski, Arianna Gabrieli, Dario Bernacchia, Stefano Rusconi, Giuliano Rizzardini, Spinello Antinori, Massimo Galli

## Abstract

This report describes the isolation, the molecular characterization and the phylogenetic analysis of the first three complete genomes of SARS-CoV-2 isolated from three patients involved in the first outbreak of COVID-19 in Lombardy, Italy. Early molecular epidemiological tracing suggests that SARS-CoV-2 was present in Italy weeks before the first reported cases of infection.

## INTRODUCTION

A new coronavirus causing severe respiratory diseases, closely related to SARS-CoV, called SARS-CoV-2, was described for the first time in the city of Wuhan, in the Hubei province of internal China, on late December 2019 (https://www.who.int/csr/don/05-january-2020-pneumonia-of-unkown-cause-china/en/). The virus belongs to the β-coronavirus genus of the Coronaviridae family showing a 96% genomic identity with a previously detected SARS-like bat coronavirus ^1,2^. Since then the virus has spread in several continents being declared, on 30 January 2020, from the World Health Organisation (WHO) a Public Health Emergency of International Concern (https://www.who.int/news-room/detail/30-01-2020-statement-on-the-second-meeting-of-the-international-health-regulations-(2005)-emergency-committee-regarding-the-outbreak-of-novel-coronavirus-(2019-nCoV). As of 13 March 2020, the now called COVID-19, caused a total of 137,445 cases and 5,088 fatalities, in 117 countries of the World (https://gisanddata.maps.arcgis.com/apps/opsdashboard/index.html#/bda7594740fd40299423467b48e9ecf6). In the last few days important concerns raised about the increasing numbers of cases outside of China, in particular in South Korea, Iran and Italy even though in February, 26 the WHO Director-General’s declared that since the day before, the number of new cases reported outside China exceeded the number of new cases in China (https://www.who.int/dg/speeches/detail/who-director-general-s-opening-remarks-at-the-mission-briefing-on-covid-19---26-february-2020).

Following the imported cases of two Chinese travellers on January 31, on February 21, the first cluster of 16 Italian cases have been reported in the Italian Northern region of Lombardy. Since than the number of notified new cases has grown exponentially, reaching, as of March, 13, a total of 15,113 cases and 1,016 deaths. Other confirmed cases of infection were then reported in several Italian regions such Veneto, Emilia-Romagna, Piemonte, Liguria and Marche.

Simultaneously, several cases suspected to be acquired in Italy, have been described in other countries out of Italy.

We have now made the isolation of the virus, the molecular characterization and the phylogenetic analysis of three complete genomes of SARS-CoV-2 isolated from three of the first 16 cases observed, none of whom reported a recent history of travel outside Italy.

## PATIENTS AND METHODS

All data used in the study were previously anonymized, according to the requirements set by Italian Data Protection Code (leg. Decree 196/2003) and by the General authorizations issued by the Data Protection Authority. Approval by Ethics Committee was deemed unnecessary because, under Italian law, such an approval is required only in the hypothesis of prospective clinical trials on medical products for clinical use (art. 6 and art. 9, leg. Decree 211/2003). Written informed consent for any other medical procedures/interventions performed for routine treatment purposes was collected for each patient.

The first subject (F.A.) was 71 years-old, he had been transferred to Sacco Hospital form the ICU of Lodi Hospital. The onset of his symptoms was February 15 and moved to our Unit on February 21, with a positive result for SARS-CoV-2 from a naso-pharingeal swab. He died on March 3 from respiratory failure. The second subject (Z.B.) was 80 years-old, he had been transferred to our Hospital form the ICU of Lodi Hospital. The onset of his symptoms was February 15 and moved to our Unit on February 22, with a positive result for SARS-CoV-2 from a naso-pharingeal swab. He died on March 1 from respiratory failure.

The third subject (R.G.) was 76 years-old, he had been transferred to our Hospital form the Internal Medicine ward of Lodi Hospital. The onset of his symptoms was February 13 and moved to our Unit on February 22, with a positive result for SARS-CoV-2 from a naso-pharingeal swab. He died on March 8 from respiratory failure.

All three subjects presented with fever (> 38.5 °C), dry cough and dyspnea on top of metabolic co-morbidities and/or hypertension. They all had a controlled mechanical ventilation and resuscitation assistance, and started treatment with lopinavir/ritonavir 400/100mg oral solution bis-in-die the same day of admission to Sacco Hospital. Their chest X-ray presented bilateral ground-glass opacities of lung bases and a certain degree of interstitial-alveolar edema, which did not improve during their hospital stay.

After isolating the virus in Vero cells, SARS-CoV-2 RNA was extracted from the culture supernatant after 24 hours, and the full genome was obtained by amplifying 26 fragments using previously published specific primers ^3^. The PCR products were used to prepare a library for Illumina deep sequencing using a Nextera XT DNA Sample Preparation and Index kit (Illumina, San Diego, California, USA) in accordance with the manufacturer’s manual, and sequencing was carried out on a Illumina MiSeq platform using the 2×150 cycle paired-end sequencing protocol. The results were mapped and aligned to the reference genome obtained from Gisaid (https://www.gisaid.org/, accession ID: EPI_ISL_412973) using Geneious software, v. 9.1.5 (http://www.geneious.com) ^4^.

The genomes obtained from the three Italian patients’ were aligned with a total of 157 SARS-CoV-2 genomes obtained worldwide and publicly available at GISAID on 3 March 2020 (https://www.gisaid.org/), and an additional Italian strain that became available during the study. Table S1 shows the accession IDs, and sampling dates and locations of the sequences included in the dataset.

In order to investigate the temporal signal of the data set, a root-to-tip regression analysis was performed by using TempEst ^5^.

The Hasegawa-Kishino-Yano model with a proportion of invariant sites (HKY+I) was selected as the simplest evolutionary model by means of JmodelTest, v. 2.1.7 (3), and the phylogenetic analysis was made using a Bayesian Markov Chain Monte Carlo (MCMC) method implemented in BEAST, v.1.8.4 ^6^.

Two coalescent priors (constant population size, exponential growth) and strict vs. relaxed molecular clock models were tested by means of Path Sampling (PS) and Stepping Stone (SS) sampling ^7^. Because the temporal signal was weak in the dataset (see Results section) the evolutionary rate was set to the values obtained from recent independent estimates^8^ (http://virological.org/t/phylodynamic-analysis-176-genomes-6-mar-2020/356). The time of the most recent common ancestor (tMRCA) were calculated in days as the unit of time.

All of the genes were tested for selection pressure using Datamonkey (https://www.datamonkey.org/).

## RESULTS

The investigation of the temporal signal in the data set by root-to-tip regression exhibited a relatively weak association between genetic distances and sampling days (correlation coefficient=0.46 and a coefficient of determination R^2^ of 0.21, Fig.1).

**Fig. 1.**
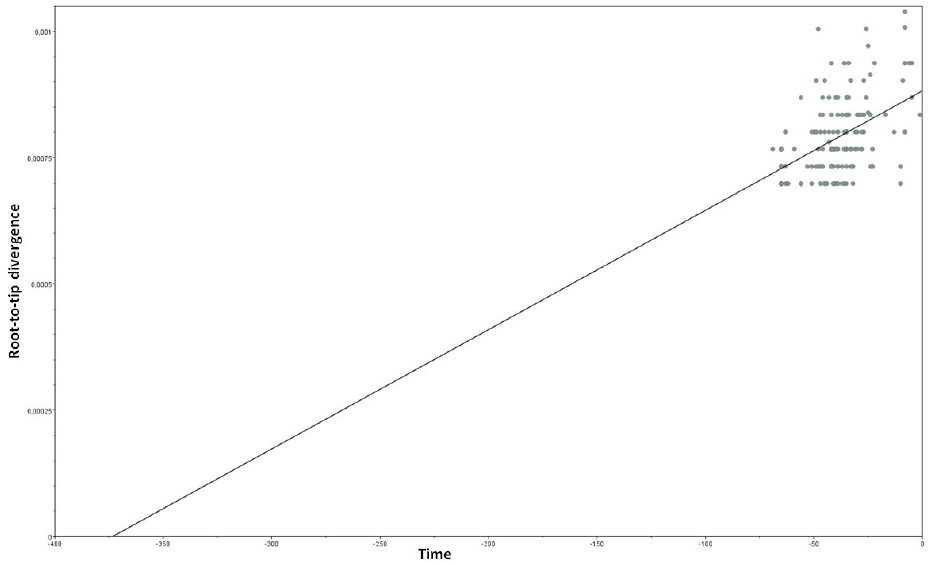
Root-to-tip regression analysis of the 161 SARS-CoV-2 sequences aligned.

The comparison of the marginal likelihoods of strict vs relaxed molecular clock and constant vs exponential coalescent models showed that the model best fitting the data was the exponential coalescent prior (PS BF exponential growth vs. constant =968.2, SS BF exponential growth vs. constant =-967.5) under a log-normal relaxed clock (PS BF strict vs. relaxed clock =-2, SS BF strict vs. relaxed clock =-1.6).

Figure 2 shows the dated tree obtained: the isolates from China were intermixed throughout the tree, mainly in a basal position with respect to the other sequences. The three Italian genomes clustered in a single highly supported clade (clade A, highlighted in Figure 1; posterior probability, pp=1) that also included two recently characterised genomes from Italy obtained from patients involved in the same outbreak in Lombardy, three isolates from Europe (two from Germany, one from Finland), and two Latin American sequences (one from Mexico and one from Brazil). One of the German isolates (from Bavaria, EPI_ISL_406862) was at the outgroup of the highly supported subclade (pp=1) including all of the other strains, and the other German sequence shared a highly supported node (pp=1) with the Mexican sequence.

**Fig. 2.**
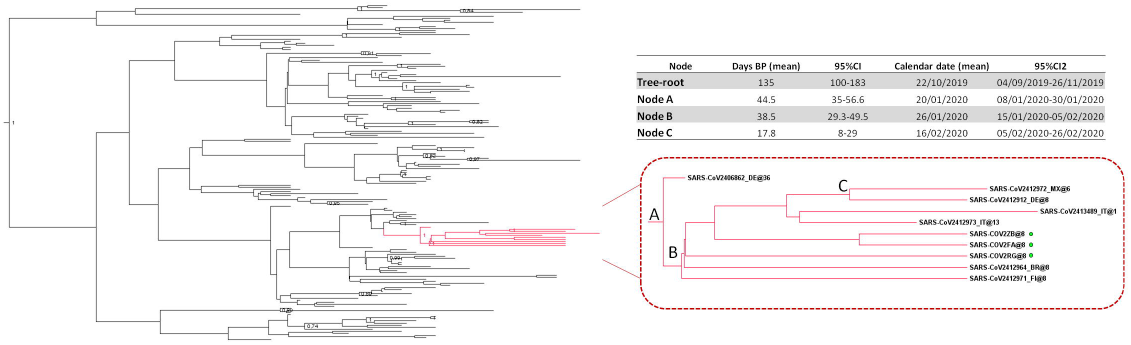
Dated tree of 161 SARS-CoV-2 showing statistically significant support for clades along the branches (posterior probability >0.7). Clade A containing the Italian strains is highlighted in red. Patients characterized in this study are evidenced with a symbol. In the table are reported tMRCA estimates and 95% HPD of the clade A significant nodes.

Table 1 summarizes the estimated mean tMRCAs of the tree root and of the main significant clades (node A and node B and node C in Figure 1). The estimated mean tMRCA of the tree-root was 135 days before the present (95%HPD: 100.4-182.6), corresponding to 23 October 2019 (credibility interval 4 September-26 November). The estimated mean tMRCA of node A was 44.5 days before the present (95%HPD: 35-56.6), corresponding to 20 January 2020 (credibility interval 8-30 January 2020), and the estimated mean tMRCA of the node B was 38.5 days before the present (95%HPD: 29.3-49.5), corresponding to 26 January 2020 (credibility interval 15 January-5 February). Finally, the estimated mean tMRCA of the internal node between the Mexican and the German isolates was 17.8 days before the present (95%HPD: 8-29), corresponding to 16 February 2020 (credibility interval 5-26 February).

Comparison of the genetic distances estimated on the basis of the number of nucleotide substitutions indicated a mean 7.8 nucleotide substitutions between the isolates in Clade A (range 0-24 nucleotide substitutions). Three sequences (two Italian and one Brazilian) were identical, whereas one Italian strain had a difference of as many as 18 nucleotides from the outgroup sequence from Germany (EPI_ISL_406862). All of the sequences in clade A showed a D614G mutation in S gene. No sites were identified as being under significant positive selection pressure.

## DISCUSSION

The present phylogenetic analysis confirms that the common origin of the SARS-CoV-2 strains characterised so far was several weeks before the first cases of COVID-19 pneumonia were described in China ^8^. It also showed that the whole genomes of the three SARS-CoV-2 strains isolated from patients in northern Italy and characterised by us are closely related to each other, as well as to the other two published Italian sequences, and the German, Finnish, Mexican and Brazilian sequences, all of which formed a highly supported clade. One of the German sequences was at the outgroup of the clade.

This sequence came from a COVID-19 outbreak reported between 20 and 24 January and occurring after business meetings with a Shanghai business woman who tested positive after returning to China ^9^. Our tMRCA estimation showed that the root of clade A was in the month of January 2020 a period compatible with this event. Nevertheless, our data cannot allow to make hypotheses on the possible routes followed by the virus to reach Italy, because it is impossible to infer the directionality of transmission, given the limited number of sparsely sampled sequences in the tree, which means that multiple independent importations to Europe cannot be excluded.

Our data suggest that SARS-CoV-2 virus entered Northern Italy between the second half of January and early February 2020, weeks before the first Italian case of COVID-19 was identified and therefore long before the current containment measures were taken.

Interestingly, although they were sampled in the same area on the same day, the genomes isolated from these three patients have a number of different, mainly synonymous substitutions. In particular, one patient living near the municipality in which the highest number of cases was recorded showed a high degree of genomic heterogeneity, thus suggesting considerable genetic drift.

In conclusion, our data show that the SARS-CoV-2 isolates which infected the Italian patients involved in the early epidemic in northern Italy and also isolated from other European and Latin America from patients reporting contacts with Italy, are closely related to the same strain isolated during one of the first European clusters, occurred in Bavaria in late January 2020 ^10^. On the basis of the phylogenetic analysis alone we cannot exclude possible multiple introductions in Germany and Italy from China (or other countries). Nevertheless, the epidemiological data showing that the first cases in Germany preceded the first ones in Italy by almost a month, suggest that the strain entered Germany before Italy. Finally, we have characterized only three genomes and we cannot exclude the presence in Italy of other different strains which could be the result of multiple introductions. Further epidemiological and molecular investigations on a larger sample are needed to clarify all these issues.

## Data Availability

A paper have been submitted to Journal of Medical Virology.

## ACKNOWLEDGEMENTS

We acknowledge the authors, originating and submitting laboratories of the sequences from GISAID.

## AUTHOR CONTRIBUTIONS

AL, GZ and MG conceived and designed the study. LM, AR, DB, SR, GR, SA and MG were involved in patients care and in the collecting biological materials. AL, AB, MT, AG and CB performed the experiments.

GZ, AL and AB performed the phylogenetic analyses. AL, GZ, AB, SR and MG wrote the first draft of the manuscript. All authors contributed to manuscript revision, read and approved the submitted version.

## DATA AVAILABILITY

Sequencing data used for the present study will be submitted to Gisaid (accession numbers to be given).

